# Parkinson’s disease progression is shaped by longitudinal changes in cerebral compensation

**DOI:** 10.1101/2025.03.21.25324393

**Authors:** Martin E. Johansson, Ivan Toni, Bastiaan R. Bloem, Rick C. Helmich

## Abstract

Parkinson’s disease (PD) is a common and debilitating neurodegenerative disorder characterized by motor slowing (bradykinesia), which is thought to arise mainly due to nigro-striatal dopaminergic cell loss. Paradoxically, longitudinal changes in striatal dopamine relate poorly to the progression of bradykinesia, indicating that other pathophysiological mechanisms play a role. In line with this, cross-sectional studies have shown that more benign motor phenotypes of PD are characterized by increased activity in the parieto-premotor cortex, indicative of cerebral compensation. However, the role of cerebral compensation in disease progression remains unclear. Here, we used a longitudinal design to test the hypothesis that the clinical progression of bradykinesia in PD is related to a decline in compensatory parieto-premotor function, over and above worsening nigro-striatal cell loss.

We used a validated action selection task in combination with functional MRI to measure motor-and selection-related brain activity in a large sample of 351 PD patients (≤5 years disease duration) and 60 healthy controls. In addition, we used diffusion-weighted MRI to obtain structural indices of substantia nigra and cerebral cortex integrity. These measurements were acquired at baseline and at two-year follow-up, enabling us to compare longitudinal changes in brain metrics between patients and controls, and to investigate their relationships with clinical metrics of bradykinesia progression.

Consistent with our hypothesis, we observed that bradykinesia progression was inversely related to longitudinal changes in selection-related dorsal premotor cortex activity, suggesting that faster loss of cortical compensation contributes to faster symptom worsening. Importantly, this relationship remained after adjusting for longitudinal changes in the functional and structural integrity of the nigro-striatal system, indicating that bradykinesia progression is uniquely determined by loss of cortical compensation. In group comparisons of longitudinal change, PD patients showed an overall reduction in putamen activity, which did not decrease further over time, in combination with an acceleration of structural decline in the substantia nigra and the premotor cortex. Despite showing expected patterns of PD pathology, neither of these metrics correlated with bradykinesia progression.

We conclude that the progression of bradykinesia in PD is determined mainly by longitudinal changes in compensatory premotor cortex function, rather than by deterioration of the nigro-striatal dopaminergic system. This presents opportunities to develop new progression-slowing interventions that focus on preserving and enhancing cortical compensation.

## Introduction

Parkinson’s disease (PD) is an increasingly common neurodegenerative disorder characterized, among other symptoms, by progressive motor slowing (bradykinesia), which gradually impairs the ability to engage in everyday activities.^1–3^ Disease-modifying treatments are urgently needed, but their development is currently hindered by a lack of biomarkers that can adequately monitor the pathophysiological mechanisms that drive the progression of symptoms.^4,5^ Converging evidence now indicates that the clinical progression of PD is partly determined by changes in compensatory hyperactivity in the cerebral cortex,^6–9^ and not just nigro-striatal dysfunction - the pathophysiological hallmark of PD - which opens possibilities to develop new biomarkers that can be leveraged to advance treatment development. Here, we exploit a unique two-year longitudinal dataset to test the hypothesis that bradykinesia progression relates more strongly to declining cortical compensation than to loss of nigro-striatal function.

The motor symptoms of PD, and bradykinesia in particular, have traditionally been attributed to loss of dopaminergic cells in the substantia nigra and depletion of dopamine in the striatum, particularly in motor territories of the posterior putamen. In turn, striatal dopamine depletion leads to excessive inhibitory output from basal ganglia nuclei, which limits activation of cortico-striatal pathways that encode movement vigor. ^10–13^ Despite providing a compelling explanation for the presence of bradykinesia, nigro-striatal cell loss may play only a limited role in determining the clinical progression of PD: striatal dopamine depletion begins several years before the onset of bradykinesia,^14,15^ is largely complete in the posterior putamen within four years after diagnosis,^16^ and is generally a poor predictor of symptom worsening.^17–22^ These observations strongly imply that the progression of bradykinesia must be shaped by additional pathophysiological mechanisms.

PD-related deficits in the nigro-striatal system develop slowly and are continuously offset by compensatory adaptations in brain function,^23–27^ particularly involving areas that remain relatively spared from neurodegeneration during early disease stages,^8,9,28^ potentially explaining why motor symptoms appear only after approximately 60% of dopamine is lost from the posterior putamen.^29^ As nigro-striatal dysfunction worsens and neurodegeneration spreads throughout the brain, compensatory adaptations gradually fail to preserve motor function and symptoms worsen.^26^ Compensatory decline therefore constitutes a promising index of clinical progression and a potential target for disease-modifying interventions.

Recent evidence suggests that compensatory decline can be measured using functional magnetic resonance imaging (fMRI).^5,7,30^ fMRI studies have consistently shown that PD is characterized by movement-related hyperactivity in parietal and premotor regions of the cerebral cortex, which may be compensatory in nature.^8,9^ In support of this conjecture, we recently demonstrated that parieto-premotor hyperactivity, measured with fMRI in combination with a visuomotor task designed to elicit PD-related compensation, is inversely related to the severity of bradykinesia.^7^ We found no relationship between bradykinesia severity and basal ganglia activity, which was consistently reduced in individuals with PD compared to healthy controls. These cross-sectional findings indicate that parieto-premotor hyperactivity helps to maintain motor performance by compensating for reduced basal ganglia activity, providing initial support for the hypothesis that motor progression is more closely linked to declining cortical compensation than to increasing nigro-striatal dysfunction (**Fig. 1A**). This hypothesis is supported further by findings from an intervention trial demonstrating that aerobic exercise attenuates the clinical progression of PD by enhancing cortical compensation.^6^

**Fig. 1.**
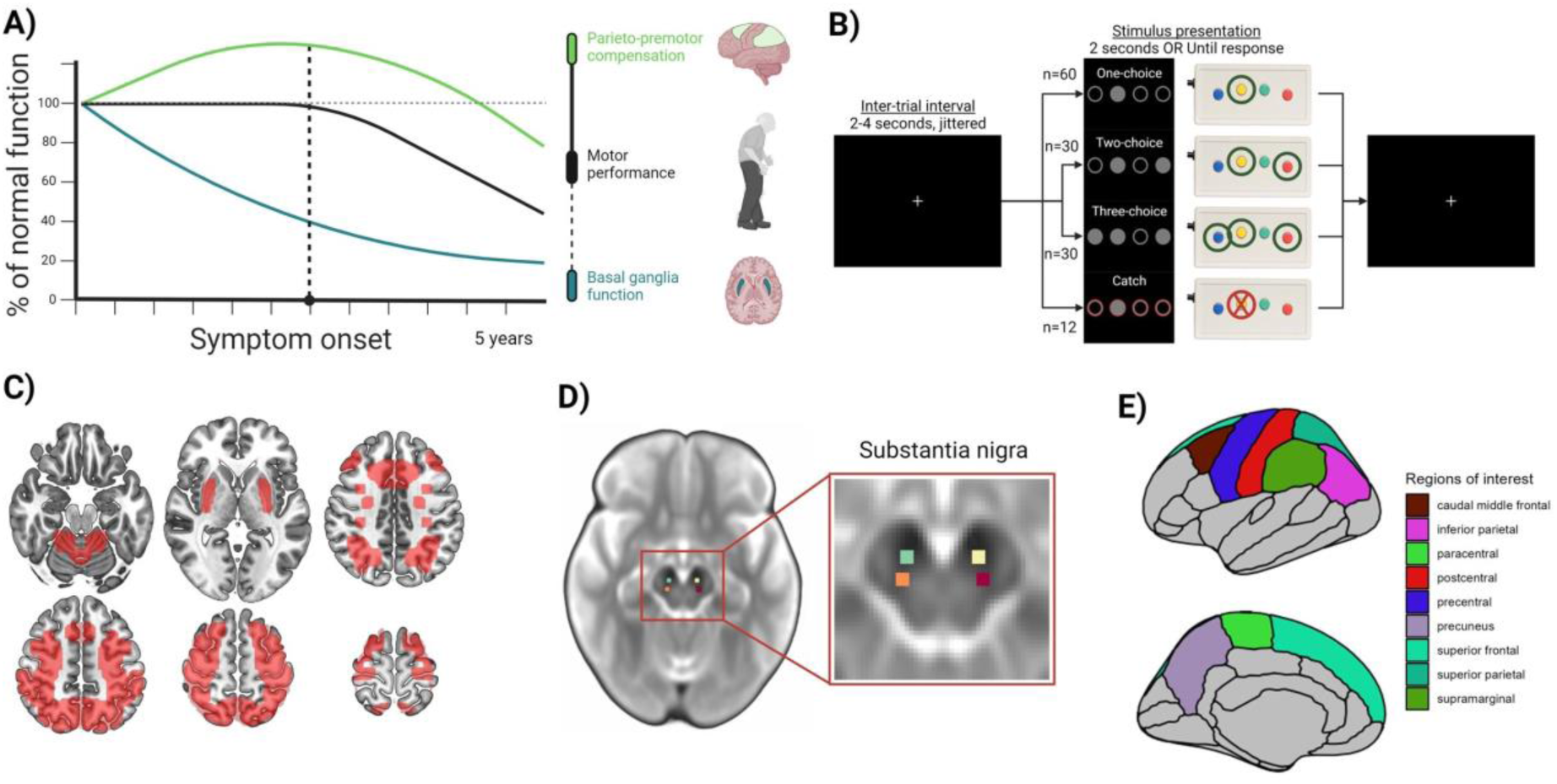
Study design. **(A)** Hypothetical model of PD progression. Loss of compensation is predicted to have a stronger influence on clinical progression than loss of basal ganglia dysfunction. Reproduced with permission from Johansson et al. 2024.^7^ **(B)** Action selection task. **(C)** Mask used to constrain voxel-wise group and brain-clinical correlation analyses to regions implicated in cortical compensation and basal ganglia dysfunction based on previous findings.^7^1 **(D)** Masks of substantia nigra sub-regions manually drawn on a study-specific b0-template and used to extract FW values. **(E)** Regions in the Desikan-Killiany atlas used to extract surface-projected MD.

Here, we combine longitudinal task-based fMRI and clinical assessments to investigate the neural mechanisms underlying individual differences in two-year clinical PD progression, focusing particularly on the role of longitudinal changes in cortical compensation. Our results indicate that the progression rates of bradykinesia are specifically shaped by loss of cortical compensation, rather than by increasing nigro-striatal dysfunction.

## Materials and methods

### Participants

367 patients diagnosed with idiopathic PD and 60 healthy controls underwent baseline MRI and clinical assessments. 338 patients and 56 healthy controls returned at two-year follow-up. 10 patients were re-diagnosed at baseline (2 parkinsonism, 8 other) and 6 patients were re-diagnosed at two-year follow-up (3 multiple-system atrophy, 2 progressive supranuclear palsy, 1 other). The exclusion of these patients led to a total sample size of 351 (baseline), and 329 (follow-up) patients. Detailed numbers of exclusions in analyses of clinical measurements, task performance, and MRI data can be found in **Supplementary Material**. All data were retrieved from the Personalized Parkinson Project (ClinicalTrials.gov identifiers: NCT03364894 and NCT05169827) database in January 2024. Written informed consent was obtained for all participants in accordance with the Declaration of Helsinki. The study was approved by a medical ethical committee (METC Oost-Nederland, formerly CMO Arnhem-Nijmegen; #2016-2934 and #2018-4785). Further details about the measurement sessions are provided in the protocol of the Personalized Parkinson Project.^31^ Demographic information can be found in **Table 1**. More detailed information about data acquisition and procedures for the estimation of task-based brain activity can be found in a previous publication.^7^

**Table 1.** Baseline characteristics and demographic information

| Measurement | Control | Patient | P-value |
| --- | --- | --- | --- |
| N (baseline/follow-up) | 60/56 | 351/328 |  |
| Age | 60.0 (9.6) | 62.7 (8.5) | 0.046 |
| Sex (F/M) | 27/33 | 129/222 | 0.283 |
| Years of education | 16.2 (3.3) | 17.2 (4.1) | 0.041 |
| Time to follow-up (years) | 2.2 (0.1) | 2.4 (0.3) | <0.001 |
| Dominant hand (L/R/None) | 55/5/0 | 295/49/7 | 0.249 |
| Responding hand (L/R) | 29/31 | 163/180 | 1.000 |
| MoCA | 27.6 (1.9) | 26.7 (2.6) | 0.004 |
| Most affected side (L/R/None) |  | 163/173/17 |  |
| Disease duration (years) |  | 2.9 (1.4) |  |
| Hoehn & Yahr-stage |  | 41/264/43/3/0 |  |
| Medication use (N/Y) |  | 17/334 |  |
| LEDD (mg) |  | 548.3 (319.8) |  |
| **MDS-UPDRS III: OFF** |  |  |  |
| Total |  | 33.5 (13.0) |  |
| Bradykinesia |  | 16.6 (7.4) |  |
| **MDS-UPDRS III: ON** |  |  |  |
| Total |  | 28.6 (12.5) |  |
| Bradykinesia |  | 14.3 (7.2) |  |

F=Female; M=Male; L=Left; R=Right; N=No; Y=Yes; mg=Milligrams; MDS-UPDRS=Movement Disorder Society Unified Parkinson's Disease Rating Scale; MoCA=Montreal Cognitive Assessment. P-values reflect outcomes of independent-samples t-tests.

### Clinical measurements

Motor symptoms were clinically assessed with part III of the Movement Disorders Society-sponsored Revision of the Unified Parkinson’s Disease Rating Scale (MDS-UPDRS III)^32^ in off-medicated (morning; <12 hours withdrawal) and on-medicated states (afternoon; approximately one hour after intake, at a time of subjective optimal improvement). Bradykinesia severity was defined as the summed scores of 11 items from the MDS-UPDRS part III (4-9 and 14).^33^ Bradykinesia progression was defined as the between-session delta (*Δ*; two-year follow-up – baseline) and was calculated for both ON- and OFF-state scores. ON-state progression was used as the primary outcome in all correlation analyses. Although sub-optimal as a marker of “pure” disease progression, ON-state progression more closely reflects the state of patients during scanning, which was done in the ON-medicated state. Furthermore, dopaminergic medication may boost compensatory mechanisms that rely on the relatively spared anterior striatum.^24^ Bradykinesia progression scores above or below three standard deviations from the mean were deemed unreliable and excluded from correlation analyses (only one patient was excluded following this criterion). Cognitive symptoms were assessed using the Montreal Cognitive Assessment (MoCA)^34^ and medication dosages were quantified as the levodopa-equivalent daily dosage (LEDD). Additionally, motor symptom asymmetry was computed as the difference between left- and right-sided bradykinetic-rigid items of the MDS-UPDRS III.^33^ Progression on these metrics were calculated the same as for bradykinesia. Missing clinical scores were imputed through predictive mean matching.^35^

### Action selection task

The behavioral task used in this study (**Fig. 1B**) and the metrics that were derived from it have been described in detail elsewhere.^7^ In short, participants were instructed to respond to a single highlighted cue with a button press as quickly and as accurately as possible, and to make equal use of all available response options. The number of highlighted cues varied from one (single-choice) to two and three (multiple-choice). Selecting between multiple, equally relevant, response options, introduces an element of mild cognitive effort that overloads the dysfunctional basal ganglia motor network in PD, thereby eliciting recruitment of compensatory resources in brain areas that remain relatively spared from pathology.^24^ The primary behavioral metric derived from the task was response times. In addition, error rates, total number of misses, response switching, and response variability were derived from the task for each participant and time point. Response times and error rates were aggregated by taking the mean across trials at each time point, trial condition, and block prior to further analysis. For response times and error rates, data were additionally summarized as motor- (mean across choices) and selection-related (multiple choice>single choice) performance at each time point to facilitate correlation analyses. Patients responded with their most affected side. The responding hand of healthy controls were matched to the patients. Time points with less than 25% correct one-choice trial responses averaged across blocks (indicating a randomized response pattern), or above 60 missing responses were deemed unreliable and therefore excluded from further analysis.

### Image acquisition and processing

#### Scanning parameters

All MRI scans were acquired using a single Siemens MAGNETOM Prisma 3T scanner (Siemens, Erlangen, Germany) equipped with a 32-channel head coil. Patients were scanned in the ON-medicated state. T1-weighted images were acquired using a magnetization-prepared rapid gradient-echo sequence (TR/TE/TI=2000/2/880 milliseconds; flip angle=8°; voxel size=1.0×1.0×1.0 millimeters; slices=192; FOV=256 millimeters; scanning time=5 minutes). Diffusion-weighted images were acquired using a multiband-accelerated echo-planar imaging sequence (TR/TE=3600/92, flip angle=90°, acceleration factor=3, scheme=monopolar, b-values=0-1000-2000/mm^2^, phase-encoding direction=A>P, voxel-size=2.0×2.0×2.0mm, slices=72, FOV=210, diffusion gradient directions=104 [5 b0 images], scanning time=6.5 minutes). Fieldmaps consisting of single b0 images with an inverted phase-encoding direction were acquired to correct for susceptibility distortions during subsequent image preprocessing. T2*-weighted images were acquired during the performance of the action selection task using a multi-band sequence (TR/TE=1000/34 milliseconds; acceleration factor=6; acquisition mode=interleaved; flip angle=60°; voxel-size=2.0×2.0×2.0 millimeters; slices=72; FOV=210 millimeters; scanning time=9-10 minutes).

### Functional MRI processing

#### Preprocessing of functional images

fMRI data were pre-processed using fMRIPrep (v23.0.2).^36^ Briefly, functional images were motion- and slice time-corrected before boundary-based registration to T1-weighted space^37^ and subsequent non-linear normalization to MNI152NLin6Asym-space (all interpolations were concatenated and applied in a single transformation).^38^ For participants with multiple time points, normalization to MNI-space was done via a participant-specific unbiased T1-weighted anatomical template.^39^ Lastly, images were smoothed with a Gaussian kernel of 6 millimeters at full-width half maximum.

#### First-level analysis

Task-related activity was estimated with first-level analyses in SPM12 (https://www.fil.ion.ucl.ac.uk/spm/software/spm12), as previously.^7^ Covariates for anatomical and motion-related sources of noise were included in each first-level model. Contrasts were set up to derive parameter estimates for two levels of choice (single choice>baseline; multiple choice>baseline), for motor-related activation (mean across choices>baseline), and for selection-related activation (multiple choice>single choice). Contrast images of left-sided responders were flipped horizontally, which ensured that the most-affected sides of patients were consistently on the right.

### Structural MRI processing

#### Preprocessing of structural images

Diffusion-weighted MRI data were pre-processed using QSIPrep (v.0.19.0).^40^ Briefly, diffusion-weighted images were first subjected to MP-PCA denoising^41^ and Gibbs unringing,^42^ followed by corrections for head motion, eddy currents, and susceptibility distortion.^43–46^ Lastly, images were resampled to 2-millimeter isotropic voxels in ACPC space. Diffusion tensor models were fitted to generate images of fractional anisotropy (FA), mean diffusivity (MD),^47,48^ and free water (FW).^49^ Additionally, average b0-images were computed. For each subject, an FA template was generated using mri_robust_register.^39^ Subject-specific FA templates from 50 healthy controls and 50 patients were combined to create a study-specific template using antsMultivariateTemplateConstruction, configured with the HCP165_FA_1mm template in MNI152NLin6Asym-space as the target.^38,50,51^

#### Substantia nigra free water

Degeneration of the pSN was estimated using a well-established FW protocol.^49^ Normalization of FW images followed previously established procedures: individual FW images were registered to subject-specific FA templates before being non-linearly normalized to the study-specific FA template.^52^ The same procedure was performed for b0 images, which were subsequently averaged and used to manually draw masks of the SN (see below).

#### Cortical surface-based mean diffusivity

Cortical degeneration was estimated as surface-based MD, a novel metric that has shown strong potential in monitoring progressive neurodegeneration,^53,54^ but which has not yet been applied to study PD. The FreeSurfer (v7.3.2) longitudinal pipeline was used to generate cortical surfaces in T1-weighted space.^39^ Boundary-based registrations^37^ (6 degrees-of-freedom) were estimated from b0-to T1-weighted space and used to project MD onto cortical surfaces.^53,54^ Per vertex, this involved averaging MD values from 6 equidistant points between the white and pial surface, starting and stopping within 20% from the borders of each surface. Using this pipeline, surface-based images were created for un-corrected and FW-corrected MD.

### Regions-of-interest

#### Functional MRI

Two regions-of-interest (ROI) were generated based on results from a previously published baseline analysis of task-related activity (**Fig. 1C**).^7^ An ROI of basal ganglia dysfunction was defined by regions showing reduced motor-related activity in patients compared to controls. This ROI included the bilateral putamen, left primary motor cortex, and right cerebellum lobule IV-V. An ROI of cortical compensation was defined by regions showing an inverse correlation with bradykinesia severity. This ROI encompassed a wide-spread network of parieto-premotor regions including the bilateral superior parietal lobule, intraparietal sulcus, superior frontal gyrus, and left middle frontal and paracingulate gyri. Bilateral versions of each ROI were constructed by adding horizontally flipped versions to the originals. Maximum filtering was used to increase spatial coverage and to merge smaller clusters into larger contiguous ones. Lastly, the two ROIs were added together to form a single, conservative ROI. This ROI was used for two purposes: to constrain voxel-wise group comparisons of longitudinal change in brain activity (i.e., investigations of the effect of TIME [baseline, follow-up]) and when correlating longitudinal changes in activity with clinical progression.

An additional ROI was constructed to investigate the reliability of previously demonstrated cross-sectional correlations between selection-related activity and off-state bradykinesia severity (**Fig. 3B, top**).^7^ This ROI was used solely for correlating selection-related activity with on-state bradykinesia, separately for baseline and two-year follow-up.

#### Structural MRI

The anterior and pSN were identified and drawn on an average b0 image, constructed from diffusion-weighted data of the same 50 healthy controls and 50 patients that contributed to the study-specific FA template (**Fig. 1D**).^52,55^ This procedure involved localizing the horizontal slice directly below the most inferior part of the red nucleus, where the SN appears hypointense. Masks of the anterior and pSN were drawn as 3×3 millimeter squares along three consecutive horizontal slices, and subsequently used to extract FW values. MD values were extracted from 9 cortical regions of the Desikan-Killiany atlas^56^ (**Fig. 1E**) in subject-specific space.

### Statistical analysis

#### Longitudinal alterations in clinical severity, behavioral performance, brain activity, and brain structure

Longitudinal changes in clinical severity and behavioral performance were investigated with linear mixed-effects modelling in R 4.2.1 (R Core Team, 2022) using the lme4-package.^57^ Models were fitted using a restricted maximum likelihood approach and *P*-values were derived from two-sided Wald *χ*^2^ tests in type III analyses of deviance. Age, sex, and years of education were modelled as covariates of non-interest in all analyses. Disease duration (i.e., years since diagnosis) and dominance of the responding hand were added as additional covariates where appropriate. Dependent variables were log-transformed whenever possible.

In analyses of bradykinesia progression, TIME (baseline, follow-up) and MEDICATION (ON, OFF) were included as within-subjects factors. Repeated measurements were accounted for with by-subject random intercepts. Individual differences in progression rates were modelled with by-subject random slopes for TIME. Longitudinal changes in MoCA scores and LEDD were analyzed similarly after omitting the MEDICATION factor and by-subject random slopes.

In comparisons of behavioral performance between patients and controls, GROUP (control, patient) was included as a between-subjects factor. TIME (baseline, follow-up) and CHOICE (single, multiple) were included as within-subjects factors. TRIAL BLOCK (one, two, three) was included as an additional within-subject factor to account for task habituation. Factor CHOICE and TRIAL BLOCK were omitted in analyses of button press variability, switching rates, and misses. Linear mixed-effects models were used to analyze response times, button press variability, and switching rates. Generalized linear mixed-effects models were used to analyze error rates (logistic regression) and misses (Poisson regression). The logistic mixed-effects model of error rates was weighted by number of trials to account for data aggregation. By-subject random intercepts accounted for repeated measurements and by-subject random slopes for TIME were included when possible.

Group comparisons in brain activity were performed with AFNI’s 3dLMEr function,^58,59^ closely following the analysis that was carried out for response times. GROUP (control, patient) was included as a between-subjects factor. TIME (baseline, follow-up) and CHOICE (single, multiple) were included as within-subjects factors. By-subject random intercepts accounted for repeated measurements. By-subject random slopes of TIME were additionally included. Comparisons involving the effect of TIME were constrained to the ROI of basal ganglia dysfunction and cortical compensation (**Fig. 1C**). Reproducibility of previously observed effects for GROUP and CHOICE were investigated separately for each session, using a whole-brain mask to avoid circularity. Reproducibility of the relationship between bradykinesia severity and selection-related activity was assessed for each sessions by extracting the average beta estimate from a network showing this relationship at baseline (**Fig 3B, top**). Cluster-forming thresholds were set at *P*<0.001 (*Z*=3.1).^60^ Multiple comparisons-corrected cluster extent thresholds were derived using 3dClustSim (10,000 iterations), based on the auto-correlation function of residuals estimated using 3dFWHMx, and considered significant at *P*<0.05 (two-sided test, *NN*=2).^61^ Anatomical labels were derived using the Anatomy Toolbox v1.8^62^ and Glasser^63^ atlases.

Group comparisons of pSN FW and cortical MD were subjected to similar linear mixed-effects modelling in R, omitting by-subject random slopes for TIME. For the pSN, an additional analysis was conducted within patients after adding HEMISPHERE (contralateral to most-versus least-affected side of the body) as a within-subjects factor. *P*-values in analyses of cortical MD were additionally adjusted for number of regions (*n*=9) using the false-discovery rate (FDR) method.

#### Associations with bradykinesia progression

Bradykinesia progression was correlated with longitudinal alterations in motor- and selection-related measurements of behavioral performance and brain activity, and with measurements of brain structure (pSN FW and cortical MD) using multiple linear regression. To ensure that these associations related specifically to intra-subject variability, and to account for regression to the mean effects, baseline scores for outcome and predictor variables were included as covariates of non-interest.^64^ Age, sex, years of education, disease duration, and dominance of the responding hand were included as additional covariates. For linear regressions between clinical and behavioral measurements, *P*-values were derived from *F*-tests in type III ANOVAs.

Four analyses were conducted with respect to bradykinesia progression in patients with complete data from both time points. First, the degree to which changes in ON- and OFF-state bradykinesia co-varied was explored. Second, change in motor- and selection-related behavioral performance was modelled as a function of bradykinesia progression. Third, associations were quantified between bradykinesia progression and brain activity change in a voxel-wise fashion using FSL’s randomise function. Here, baseline brain activity was modelled as a voxel-wise covariate and non-parametric permutation testing,^65^ combined with threshold-free cluster enhancement,^66^ was used to derive multiple-comparisons corrected cluster extent thresholds. Fourth, associations with bradykinesia progression for pSN FW and cortical MD were quantified the same as for measurements of behavioral performance. pSN FW was additionally related to motor symptom asymmetry.^10^ Analyses of bradykinesia progression accounted for longitudinal changes in cognitive performance and medication changes to enhance inference specificity. Control analyses were conducted to reproduce previous findings by correlating on-state bradykinesia severity with behavioral and brain measurements separately for baseline and follow-up.

## Results

### Clinical progression

#### Bradykinesia worsens over time, independently of medication status

Bradykinesia increased by approximately three points over two years (**Fig. 2A**; TIME effect: *χ*^2^(1)=84.01, *P*<0.001, *η^2^_p_*=0.21; follow-up>baseline: *β*=3.32, *SE*=0.36, *t-ratio*(321)=9.17, *P*<0.001) and it decreased following medication (linear mixed-effects model, MEDICATION effect: *χ*^2^(1)=282.00, *P*<0.001, *η^2^_p_*=0.31; on>off: *β*=-2.78, *SE*=0.17, *t-ratio*(636)=16.79, *P*<0.001). There were no differences in bradykinesia progression between on- and off-medicated states (TIME×MEDICATION effect: *P*=0.14). Bradykinesia progression in on- and off-medicated states correlated substantially (multiple linear regression: *F*(1)=395.83, *P*<0.001, *η^2^_p_*=0.64). Given the strong correspondence between off- and on-state clinical scores, only the latter was used for subsequent analyses.

**Fig. 2.**
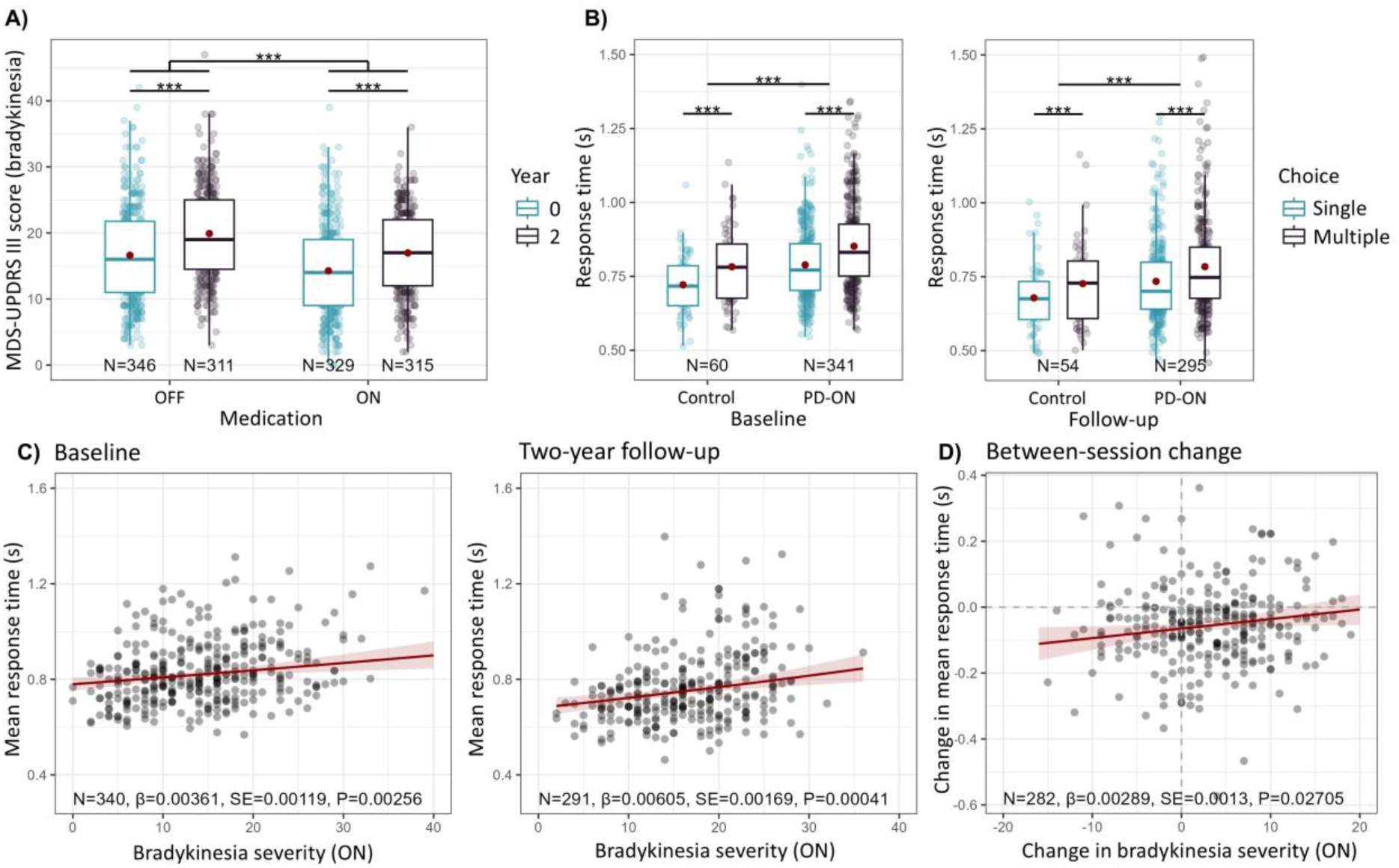
Clinical progression and longitudinal alterations in behavioral performance. **(A)** Two-year bradykinesia progression. **(B)** Response times in the action selection task. **(C)** Correlation between bradykinesia progression and longitudinal change in mean response times **(D)** Correlations between ON-state bradykinesia severity and mean response times at baseline (**left**), follow-up (**right**). ***=P<0.001.

#### Bradykinesia progression co-varies with cognitive decline and medication titration

Over time, cognitive performance declined (TIME effect: *χ*^2^(1)=54.26, *P*<0.001, *η*^2^ =0.15; follow-up>baseline: *β*=-0.41 [equivalent to a 1.12 point decline in non-adjusted MoCA scores], *SE*=0.06, *t-ratio*(301)=7.37, *P*<0.001) whereas medication dosages increased (TIME effect: *χ*^2^(1)=165.40, *P*<0.001, *η^2^_p_*=0.35; follow-up>baseline: *β*=217.03, *SE*=16.88, *t-ratio*(306)=12.86, *P*<0.001). Furthermore, longitudinal decline in cognitive performance correlated with bradykinesia progression (multiple linear regression: *F*(1)=13.80, *P*<0.001, *η*^2^ =0.002, *β*=-0.03, *SE*=0.01), although increasing medication dosages did not (multiple linear regression: *P*=0.45). To enhance the specificity of associations involving bradykinesia, cognitive decline and medication titration were added as covariates of non-interest in subsequent correlation analyses.

### Longitudinal analyses of behavioral performance

#### Parkinson’s disease impairs overall motor performance

Patients responded slower compared to controls (**Fig. 2B**; GROUP effect: *χ*^2^(1)=11.18, *P*<0.001, *η*^2^ =0.03; patient>control: *log-ratio*=1.07, *SE*=0.021, *t-ratio*(370)=3.34, *P*<0.001). Additionally, all participants responded slower as demands on action selection increased, although this was less pronounced at follow-up compared to baseline (linear mixed-effects model, TIME×CHOICE: *χ*^2^(1)=4.67, *P*=0.031, *η*^2^ =0.001; follow-up>baseline, multiple>single: *log-ratio*=0.99, *SE*=0.006, *t-ratio*(4941)=2.16, *P*=0.031). There was no indication that PD patients became slower over time compared to healthy controls (linear mixed-effects model, GROUP×TIME and GROUP×TIME×CHOICE effects: both *P*>0.69).

Analyses of additional behavioral metrics (error rates, number of misses, response flexibility) showed that PD patients and healthy controls performed the action selection task using similar strategies (**Supplementary Material**).

#### Behavioral performance indexes bradykinesia progression and severity

Faster bradykinesia progression was associated with longitudinal slowing of motor-related response times (**Fig. 2C**; *F*(1)=4.94, *P*=0.027, *β*=2.893e-03, *SE*=1.302e-03). Additionally, motor-related response times increased as a function of on-state bradykinesia severity at both baseline (**Fig. 2D**; multiple linear regression: *F*(1)=9.24, *P*=2.561e-03, *β*=3.609e-03, *SE*=1.187e-03) and follow-up (multiple linear regression: *F*(1)=12.81, *P*=4.068e-04, *β*=6.046e-03, *SE*=1.690e-03). Error rates and number of misses correlated similarly with bradykinesia severity (**Supplementary Material**).

### Longitudinal analyses of task-related brain activity

#### Parkinson’s disease impairs movement-related processing in the basal ganglia

PD patients showed reduced motor-related activity in the putamen compared to healthy controls (**Fig. 3A, top**; **Table 2**). This effect was present at the whole-brain level at both baseline and at follow-up. Additionally, increasing action selection demand reliably elicited strong parieto-prefrontal network activation and default-mode network deactivation for both PD patients and healthy controls (**Fig. 3A, bottom**). However, analogous to the analysis of response times, there were no differences in longitudinal change of selection-related activity between PD patients and healthy controls.

**Fig. 3.**
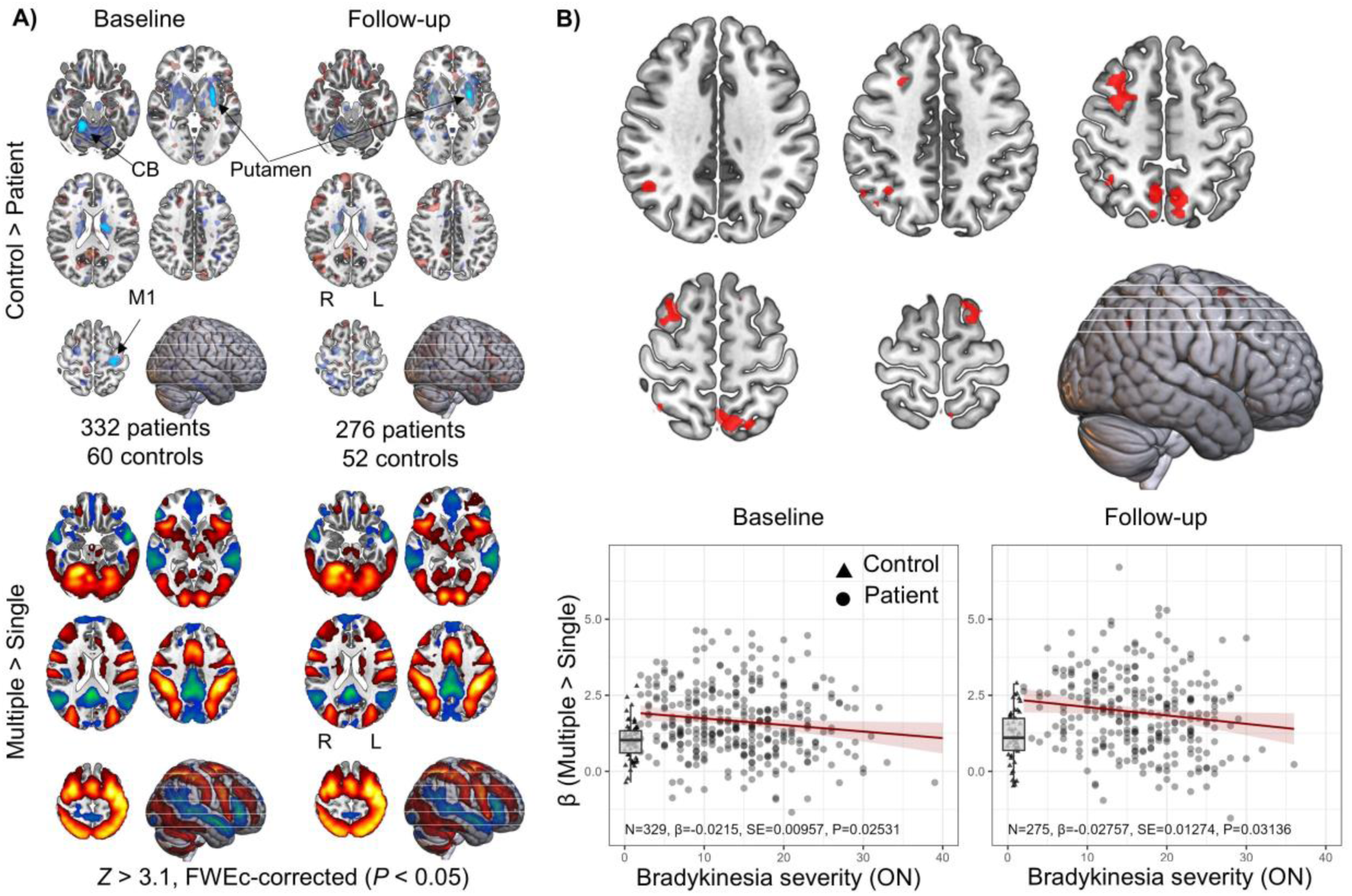
Repeatability of task-related effects and brain-clinical correlations. **(A)** Reduced movement-related putamen activity in patients compared to controls (top) and the effect of increasing action selection demand (bottom) is present at both baseline and two-year follow-up. **(B)** Correlations between selection-related activity and off-state bradykinesia severity observed at baseline in a previous publication (top) is also present in the on-state at baseline (bottom-left) and at follow-up (bottom right). Boxplots display the range of selection-related activity for healthy controls. FWEc=Familywise error-corrected, cluster-level; CB=Cerebellum; M1=Primary motor cortex; R=Right; L=Left; SE=Standard error of the mean.

**Table 2.** Voxel-wise mixed-effects comparisons of longitudinal changes in brain activity between patients and healthy controls

| Anatomical label (% cluster volume in area) | Area | P-value (FWEc-corrected) | Cluster extent (voxels) | FWEc-corrected cluster extent threshold ($\alpha=0.05$; $P<0.001$; voxels)<sup>2</sup> | Max z-value | MNI: X, Y, Z |
| --- | --- | --- | --- | --- | --- | --- |
| **GROUP: Control > Patient (across sessions)<sup>1</sup>** |  |  |  |  |  |  |
| L putamen (82%) |  | P<0.0001 | 650 | 36 | 7.65 | -26,-2,3 |
| R putamen (67%) |  | P<0.0001 | 183 | 36 | 5.75 | 28,-6,0 |
| R cerebellum (88%) | IV-V | P<0.0001 | 135 | 36 | 4.84 | 16,-50,-20 |
| L precentral gyrus (94%) | MI | P<0.02 | 51 | 36 | 4.07 | -28,-24,63 |
| **GROUP: Patient > Control (across sessions)<sup>1</sup>** |  |  |  |  |  |  |
| Ns. |  |  |  | 36 |  |  |
| **GROUP: Control > Patient (baseline)<sup>1</sup>** |  |  |  |  |  |  |
| L putamen (82%) |  | P<0.0001 | 470 | 36 | 7.26 | -27,-4,3 |
| R cerebellum (92%) | IV-V | P<0.0001 | 171 | 36 | 6.23 | 16,-48,-20 |
| L precentral gyrus (92%) | MI | P<0.0001 | 131 | 36 | 4.61 | -29,-25,62 |
| R putamen (96%) |  | P<0.02 | 52 | 36 | 5.14 | 30,-5,4 |
| **GROUP: Patient > Controls (baseline)<sup>1</sup>** |  |  |  |  |  |  |
| Ns. |  |  |  | 36 |  |  |
| **GROUP: Control > Patient (follow-up)** |  |  |  |  |  |  |
| L putamen (91%) |  | P<0.0001 | 189 | 36 | 4.74 | -26,-1,2 |
| R putamen (54%) |  | P<0.03 | 46 | 36 | 4.58 | 28,-9,1 |
| **GROUP: Patient > Controls (follow-up)** |  |  |  |  |  |  |
| Ns. |  |  |  | 36 |  |  |
| **GROUP×TIME** |  |  |  |  |  |  |
| Ns. |  |  |  | 36 |  |  |
| **GROUP×TIME×CHOICE** |  |  |  |  |  |  |
| Ns. |  |  |  | 36 |  |  |

<sup>1</sup>Effects derived using a whole-brain mask instead of the ROI in Figure 1C, which was partially based on findings at baseline. <sup>2</sup>Simulated based on the auto-correlation function of residuals. Anatomical labels and areas are derived from the Anatomy Toolbox v1.8 (CA\_ML\_18\_MNI) and Glasser (MNI\_Glasser\_HCP\_v1.0) atlases, respectively.

#### Bradykinesia progression is determined by longitudinal changes in cortical compensation

In line with the main hypothesis, bradykinesia progression correlated negatively with longitudinal change in selection-related activity in the right and left dorsal premotor cortex (**Fig. 4**; **Table 3**). To control for potential contributions of nigro-striatal deficits, the analysis above was repeated after including longitudinal changes in selection-related putamen activity and pSN FW as additional covariates of non-interest. The previously identified cluster in the right premotor cortex remained significant, while the relatively weaker cluster in the left premotor cortex did not (**Fig. 4**; **Table 3**).

**Fig. 4.**
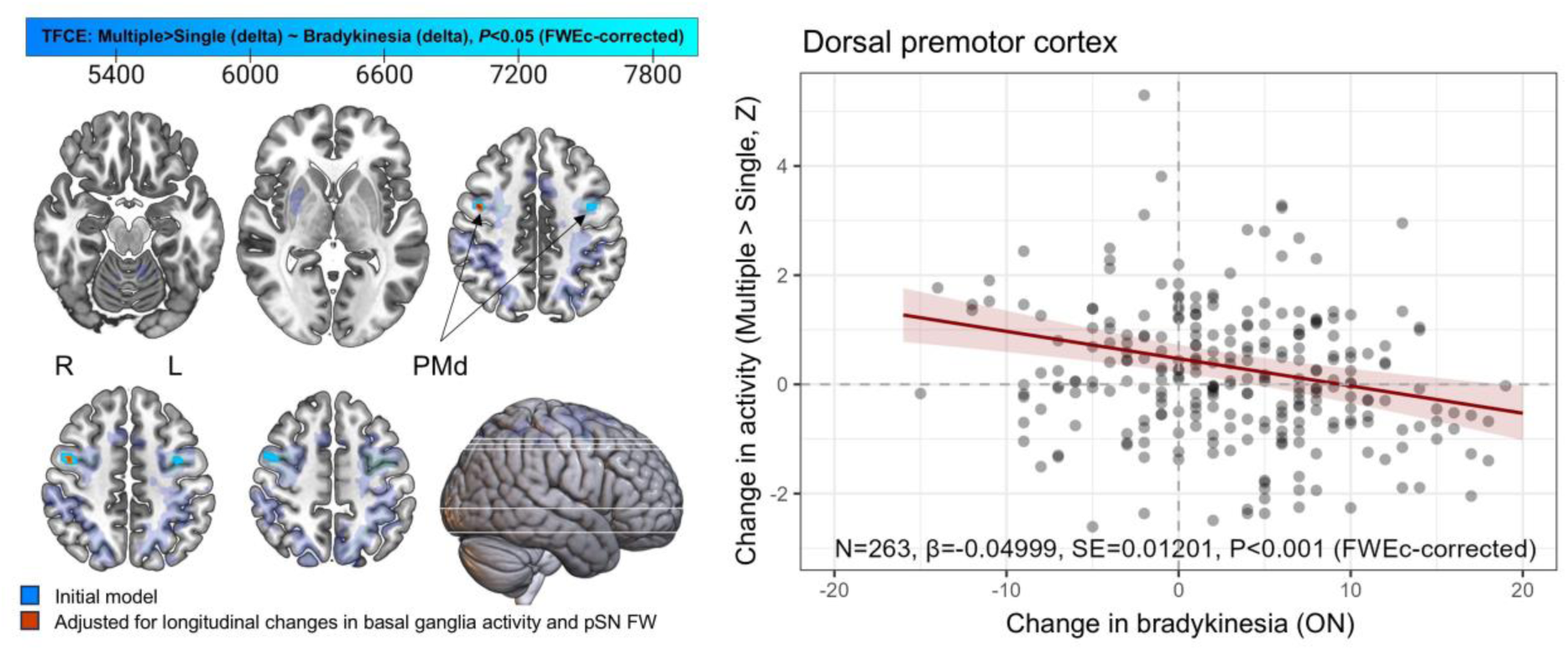
Longitudinal correlation between bradykinesia and selection-related premotor cortex activity. Two-year bradykinesia progression correlates inversely with longitudinal changes in selection-related activity across the extent of the ROI (blue), reaching the threshold for statistical significance only in the bilateral dorsal premotor cortex. After adjusting for longitudinal changes in the functional and structural integrity of the nigro-striatal system, the cluster in the right hemisphere remains significant (red). FW=Free water; FWEc=Familywise error, cluster-level; PMd=Premotor cortex, dorsal; pSN=posterior substantia nigra.

**Table 3.** Voxel-wise correlation analyses between bradykinesia progression and longitudinal changes in brain activity

| Anatomical label (% cluster volume in area) | Area | P-value (FWEC-corrected) | Cluster extent (voxels) | Max TFCE | MNI: X, Y, Z |
| --- | --- | --- | --- | --- | --- |
| ***ΔMultiple > Single (selection-related activity) ~ ΔBradykinesia*** |  |  |  |  |  |
| **Positive correlation** |  |  |  |  |  |
| Ns. |  |  |  |  |  |
| **Negative correlation** |  |  |  |  |  |
| R precentral gyrus (99%) | FEF | 0.005 | 97 | 8007 | 45,-3,44 |
| L precentral gyrus (86%) | FEF | 0.03 | 46 | 5207 | -44,-3,42 |
| ***ΔMultiple > Single (selection-related activity) ~ ΔBradykinesia, adjusted for nigro-striatal dysfunction*** |  |  |  |  |  |
| **Positive correlation** |  |  |  |  |  |
| Ns. |  |  |  |  |  |
| **Negative correlation** |  |  |  |  |  |
| R precentral gyrus (100%) | FEF | 0.022 | 28 | 5956 | 43,-3,44 |
| ***ΔMean > Baseline (motor-related activity) ~ ΔBradykinesia*** |  |  |  |  |  |
| **Positive correlation** |  |  |  |  |  |
| Ns. |  |  |  |  |  |
| **Negative correlation** |  |  |  |  |  |
| Ns. |  |  |  |  |  |

Anatomical labels and areas are derived from the Anatomy Toolbox v1.8 (CA\_ML\_I8\_MNI) and Glasser (MNI\_Glasser\_HCP\_v1.0) atlases, respectively.

#### Selection-related activity reliably indexes bradykinesia severity

In an ROI analysis based on previous cross-sectional findings,^7^ selection-related activity decreased as a function of on-state bradykinesia severity at both baseline (**Fig. 3B, lower left**; multiple linear regression: *F*(1)=5.050, *P*=2.531e-02, *η^2^_p_*=0.021) and at follow-up (**Fig. 3B, lower right**; multiple linear regression: *F*(1)=4.683, *P*=3.136e-02, *η^2^_p_*=0.034).

### Longitudinal analyses of brain structure

#### Parkinson’s disease-specific degeneration of the posterior substantia nigra

There was a trend towards a larger increase in pSN FW in PD patients compared to healthy controls (**Fig. 5A**; **Supplementary Table 1**; GROUP×TIME effect: *χ*^2^(1)=2.95, *P*=0.086). Post hoc analyses of longitudinal FW changes in each group revealed a clear increase in PD patients (follow-up>baseline: *log-ratio*=1.027, *SE*=0.008, *t-ratio*(340)=3.35, *P*<0.001), but not in healthy controls (*P*=0.60).

**Fig. 5.**
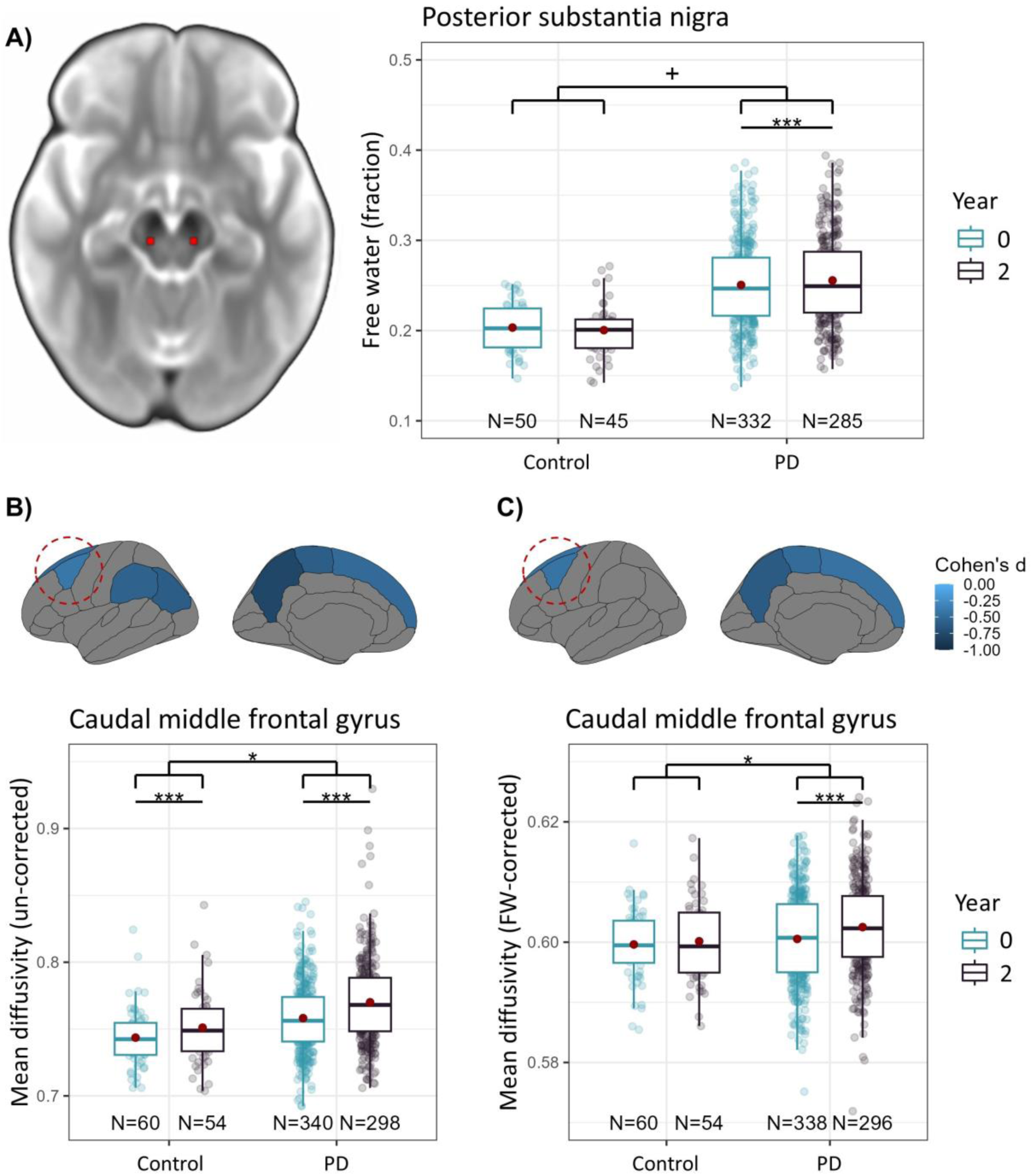
Effects of PD on longitudinal alterations in brain structure. **(A)** PD is associated with increased pSN FW **(B)**, and with increased MD in caudal middle frontal, superior frontal, inferior parietal, supramarginal, precuneus, and paracentral cortex. **(C)** After correcting for FW, increased MD is confined to the caudal middle frontal, paracentral, precuneus, supramarginal, and superior frontal cortex. +=0.086, *<0.05, ***=P<0.001.

The analysis above was repeated within PD patients after identifying the sides of the SN that were contralateral to the clinically most- and least-affected sides of the body. As before, there was an increase in pSN FW over time (TIME effect: *χ*^2^(1)=13.17, *P*<0.001; follow-up>baseline: *log-ratio*=1.048, *SE*=0.008, *t-ratio*(319)=3.63, *P*=<0.001). Additionally, FW was higher in the pSN contralateral to the clinically most-affected side of the body (HEMISPHERE effect: *χ*^2^(1)=11.38, *P*<0.001, most>least: *log-ratio*=1.048, *SE*=0.015, *t-ratio*(563)=3.37, *P*<0.001). There were no differences in longitudinal change between hemispheres (TIME× HEMISPHERE effect: *P*=0.82).

#### Posterior substantia nigra degeneration indexes motor symptom asymmetry, but not clinical progression

There was no correlation between bradykinesia progression and longitudinal changes in pSN FW (**Supplementary Table 2**; multiple linear regression: *F*(1)=0.25, *P*=0.62), nor between pSN FW and bradykinesia severity at either baseline (**Supplementary Table 3**) or follow-up (**Supplementary Table 4**).

Right-left asymmetry of motor symptoms correlated negatively with right-left asymmetry in pSN FW at both baseline (multiple linear regression: *F*(1)=9.49, *P*=0.002, *β*=-0.068, *SE*=0.022) and follow-up (multiple linear regression: *F*(1)=8.60, *P*=0.004, *β*=-0.092, *SE*=0.031). However, there was no relationship between longitudinal change in the two variables (*P*=0.64).

#### Parkinson’s disease-specific structural changes in the cerebral cortex

In 6 out of 9 cortical ROIs, PD patients showed greater longitudinal increases in cortical MD compared to controls (**Fig. 5B**; **Supplementary Table 1**). This included the caudal middle frontal cortex (GROUP×TIME effect: *χ*^2^(1)=5.54, *P_FDR_*=0.028, *η^2^_p_*=0.015) and superior frontal cortex (GROUP×TIME effect: *χ*^2^(1)=8.36, *P_FDR_*=0.007, *η*^2^ =0.023), where bradykinesia progression correlated with selection-related activity.

#### Cortical structure does not index clinical progression in Parkinson’s disease

There was no correlation between longitudinal changes in cortical MD and bradykinesia progression (**Supplementary Table 2**). At baseline, bradykinesia severity correlated with MD in the inferior parietal, precuneus, superior frontal, and supramarginal cortices (**Supplementary Table 3**). At follow-up, there were no correlations with bradykinesia severity (**Supplementary Table 4**).

To enhance the specificity of MD with respect to microstructural damage, a post-hoc analysis of FW-corrected cortical MD was done.^53^ Here, longitudinal PD-related increases in FW-corrected cortical MD compared to healthy controls were confined to 4 out of 9 cortical ROIs (**Fig. 5C**; **Supplementary Table 1**), including the caudal middle frontal cortex (GROUP×TIME effect: *χ*^2^(1)=6.72, *P_FDR_*=0.025, *η*^2^ =0.019) and the superior frontal cortex (GROUP×TIME effect: *χ*^2^(1)=6.44, *P_FDR_*=0.025, *η*^2^ =0.018). As previously, there were no correlations between FW-corrected cortical MD and bradykinesia progression (**Supplementary Table 2**). Moreover, FW-corrected cortical MD did not correlate with bradykinesia severity at either baseline (**Supplementary Table 3**) or follow-up (**Supplementary Table 4**).

#### Loss of functional compensation is not associated with localized changes in premotor cortex structure

A post hoc analysis was done to test whether loss of cortical compensation may have resulted from progressive microstructural damage. Selection-related premotor cortex activity did not correlate with surface-based MD (all *P*>0.5) or FW-corrected MD (all *P*>0.1) in either the middle or superior frontal cortex.

## Discussion

We tested the hypothesis that progressive worsening of motor symptoms in PD over two years is associated with gradual loss of compensatory function in the cerebral cortex (quantified with task-based fMRI), over and above nigro-striatal degeneration (quantified with free water imaging of the pSN).^7^ There are two main sets of findings. First, we provide empirical evidence for our hypothesis that (hyper)activity in the parieto-premotor cortex serves a compensatory role, and that the degree of cortical compensation shapes the clinical course of PD. Specifically, in line with our previous cross-sectional work, we demonstrate that bradykinesia severity correlates negatively with selection-related activity in the parieto-premotor cortex at two separate time points, further supporting the view that enhanced cortical function compensates for underlying deficits in the nigro-striatal motor network.^7^ In addition, we show that individual rates of bradykinesia progression correlate specifically with longitudinal changes in selection-related activity in the premotor cortex, even when accounting for the functional and structural integrity of the nigro-striatal system. Second, we provide empirical evidence for the canonical finding that nigro-striatal cell loss in PD leads to severe striatal dysfunction.^8–10^ Specifically, PD patients showed increased levels of pSN FW (a measure of nigral cell damage) compared to healthy controls, which progressed over two years and was more pronounced in the most-versus least-affected hemisphere. Furthermore, PD patients showed consistent reductions in motor-related activity in the putamen across two time points. However, neither of these measures correlated with bradykinesia severity or progression. Taken together, our findings indicate that the progression of bradykinesia is uniquely shaped by the loss of cortical compensation, and not just by deficits in the nigro-striatal system.

### The role of the nigro-striatal system in Parkinson’s disease progression

Despite the prominent role assigned to nigro-striatal deficits in producing the symptoms of PD,^10^ we observed no relationships between structural or functional indices of nigro-striatal integrity and clinical symptom progression. In PD patients compared to healthy controls, our imaging analyses revealed decreased putamen activity, which remained stable over two years, and elevated FW in the pSN, which increased over time. These results are consistent with previous findings from both cross-sectional and longitudinal MRI studies of PD,^8,9,55^ which supports the soundness of our methodological approach.

The previously observed lack of relationships between dopamine depletion and clinical progression may reflect how pathology propagates through the nigro-striatal system. It has been argued that the highly branched dopaminergic (and noradrenergic) neurotransmitter systems undergo a “dying-back” process of degeneration in PD, such that striatal dopamine terminals perish first, followed by loss of cell bodies in the SNpc.^67–71^ Our findings are consistent with this idea: deficits in motor-related activity in the putamen of PD patients remained unchanged over the course of two years, indicating a potential flooring effect resulting from the culmination of dopamine terminal loss,^16,22,72^ while the reduced structural integrity of the pSN deteriorated further, which suggests propagation from axon to cell body. It should be noted that we measured task-based activity while patients were on their usual dopaminergic treatment, which was continuously titrated from baseline to follow-up. Increasing medication dosages may have led to normalization of putamen activity, thereby explaining the lack of progression. Arguing against this, we have previously shown, using an on-versus-off design, that dopaminergic medication did not influence putamen activity elicited by the action selection task.^7^ Furthermore, inclusion of dopaminergic medication dose as a covariate did not change any of the findings.

Lastly, we would like to emphasize that our findings by no means suggest that nigro-striatal deficits are clinically irrelevant for PD progression: the most-affected side of the pSN reliably reflected the clinically most-affected side of the body, as did motor-related activity reductions in the putamen. Moreover, the level of asymmetric degeneration in the pSN was proportional to the asymmetric distribution of motor symptoms. This indicates that nigro-striatal deficits are a necessary condition for the development of motor symptoms (and explains why dopamine replacement therapy remains an effective treatment for alleviating motor disability), but likely without being the primary source of their progressive worsening in the years following the diagnosis.

### The role of cortical compensation in Parkinson’s disease progression

In line with our main hypothesis, we found that the progressive worsening of motor symptoms correlated with a progressive decline in dorsal premotor hyperactivity, which we argue is compensatory in nature.^7^ Hyperactivity in the cerebral cortex has been observed consistently in neuroimaging studies of PD,^8,9^ particularly in response to cognitively demanding tasks,^73^ and has often been interpreted as a compensatory phenomenon.^25^ However, hyperactivity may also reflect maladaptive changes, such as loss of inhibition, that exacerbate symptoms and contribute to disease progression.^74^ For instance while increased cortical input to the striatum may reflect a compensatory adaptation in early stages of PD, it could potentially also hasten the degeneration of nigro-striatal terminals due to increased energy demands.^71,75^ The inverse correlation between premotor cortex activity and symptom severity, both cross-sectionally at each time point and when assessing longitudinal changes, suggests that the premotor activity we observed here reflects a compensatory mechanism. A contributing factor may be that the dorsal premotor cortex is functionally connected to anterior territories of the striatum,^76^ which remain relatively spared from pathology until later disease stages.^24^ Furthermore, parietal and premotor areas are among the last cortical regions to atrophy due to synuclein pathology, potentially enabling them to retain their capacity for compensatory adjustments into later disease stages, where their functionality starts to degrade due to spreading neurodegeneration, without becoming maladaptive.^28,77^

To understand how cortical compensation can be preserved, it is essential to identify underlying processes that contribute to its decline. To this end, we used diffusion-based imaging to measure microstructural damage longitudinally in cortical grey matter.^53,54^ We demonstrate that PD leads to an accelerated structural decline in several cortical regions, including the premotor cortex. However, structural cortical changes did not correlate with the speed of clinical progression nor with changes in task-related activity. Taken together, this indicates that progressive changes in functional cortical compensation may not be directly related to localized changes in cortical structure.

Our finding that the loss of compensatory parieto-premotor cortex function relates to the speed at which bradykinesia worsens opens new opportunities to design progression-slowing interventions. During movement planning, the parieto-premotor cortex exerts causal influences over activity in the primary cortex, biasing it towards a contextually appropriate motor plan.^78^ Enhancing these biasing influences onto the primary motor cortex may enable the parieto-premotor cortex to more effectively overcome deficits in basal ganglia output. Interestingly, studies in healthy individuals have demonstrated that stimulation of the parieto-premotor cortex can alter the excitability of the primary motor cortex,^79,80^ which may translate into beneficial effects on motor performance in PD.^81,82^ Recently, it has been shown that the primary motor cortex of the clinically least-affected hemisphere compensates for loss of dopamine through enhanced plasticity and reduced interhemispheric inhibition, potentially making it more receptive to additional compensatory input from upstream cortical areas such as the premotor cortex.^83,84^ Further research is needed to ascertain how stimulation of the parieto-premotor cortex alters compensatory influences over primary motor cortex activity, and whether such influences can be leveraged to slow down clinical progression.

### Interpretational issues

The relationship we observed between individual differences in the speed of clinical progression and compensatory collapse needs to be viewed in the light of our group-level comparisons. While symptoms progressively worsened across PD patients, there was no consistent reduction in compensation over time, arguing against the possibility that its relationship with clinical progression was mediated by ageing-related influences, which occurred similarly for both patients and controls. More importantly, lack of group-level effects on compensation strongly implies that PD patients follow their own unique trajectories of compensatory collapse, rather than a strictly linear decline. These trajectories may reflect underlying heterogeneity in PD-related pathology, giving rise to subtypes characterized by opposing directions of longitudinal change that contribute to clinical progression despite cancelling out effects at a group level.^7,33,85,86^ This heterogeneity is apparent in **Fig. 3B**, where the between-subject variability of selection-related activity is markedly more pronounced for PD patients than it is for healthy controls. Detecting individual differences in trajectories of compensatory collapse may be key to identifying patients at risk of rapid progression.

It may be argued that scanning in an on-medicated state may have led to a loss in sensitivity with respect to detecting associations with “pure” disease progression. However, even after medication withdrawal, there are long-lasting effects of dopaminergic medication that provide symptomatic relief lasting for days.^87^ Moreover, understanding the mechanisms underlying disease progression in an on-medicated state holds intrinsic value as it more closely reflects the everyday life of patients.

## Conclusion

We provide empirical evidence supporting the hypothesis that clinical progression in PD is associated with a collapse of compensatory function in the dorsal premotor cortex. Further research is now required to demonstrate whether delaying this collapse can have disease-modifying effects. Emerging evidence already indicates that compensation can be preserved through life-style alterations that promote neuronal health, such as physical activity,^6,88,89^ and through targeted neuromodulation techniques that temporarily boost cortical function, which can now be applied with high precision in humans owing to recent advancements in transcranial ultrasound stimulation.^82,90^ In combination, the complex neurophysiological effects that these non-pharmacological interventions evoke may have the potential to both elicit and retain compensatory enhancements, thereby contributing to the attenuation of clinical progression in PD beyond what is currently achievable through more traditional pharmacological treatments.

## Data availability

Data supporting the findings of this study are available upon request only, to ensure the privacy of participants. A data acquisition request can be sent to the corresponding author or using the following website: https://www.personalizedparkinsonproject.com/home/data/requesting.

## Acknowledgements

We thank everyone who participated in the Personalized Parkinson Project for their contributions.

## Funding

This study was supported by The Michael J. Fox Foundation for Parkinson’s Research (grant ID #15581). The Personalized Parkinson Project was co-funded by Verily Life Sciences LLC, the city of Nijmegen and the Province of Gelderland, Radboud University Medical Center, and Radboud University. Allowance made available by Health ∼ Holland, Top Sector Life Sciences and Health, to stimulate public-private partnerships. The Centre of Expertise for Parkinson & Movement Disorders was supported by a Centre of Excellence grant from the Parkinson’s Foundation.

## Competing interests

M.E.J has no conflicts of interest to declare. I.T. has no conflicts of interest to declare. B.R.B. serves as the co-editor in chief for the Journal of Parkinson’s Disease; serves on the editorial board of Practical Neurology and Digital Biomarkers; has received fees for serving on the scientific advisory board for the Critical Path Institute, Gyenno Science, MedRhythms, UCB, Kyowa Kirin, and Zambon (paid to the institute); has received fees for speaking at conferences from AbbVie, Bial, Biogen, GE Healthcare, Oruen, Roche, UCB, and Zambon (paid to the institute); and has received research support from Biogen, Cure Parkinson’s, Davis Phinney Foundation, Edmond J. Safra Foundation, Fred Foundation, Gatsby Foundation, Hersenstichting Nederland, Horizon 2020, IRLAB Therapeutics, Maag Lever Darm Stichting, The Michael J. Fox Foundation, Ministry of Agriculture, Ministry of Economic Affairs and Climate Policy, Ministry of Health, Welfare and Sport, Netherlands Organization for Scientific Research (ZonMw), Not Impossible, Parkinson Vereniging, Parkinson’s Foundation, Parkinson’s UK, Stichting Alkemade-Keuls, Stichting Parkinson NL, Stichting Woelse Waard, Topsector Life Sciences and Health, UCB, Verily Life Sciences, Roche, and Zambon. B.R.B. does not hold any stocks or stock options with any companies that are connected to PD or to any of his clinical or research activities. R.C.H has no conflicts of interest to declare.

## Supplementary material

### Missingness of data

At baseline, 351 patients and 60 controls participated. 329 patients and 56 controls returned for two-year follow-up measurements.

#### Clinical data

At baseline, 23 ON-state (*N*=329) and 6 OFF-state (*N*=346) patients lacked clinical data. At two-year follow-up, 14 ON-state (*N*=315) and 18 OFF-state (*N*=311) patients lacked clinical data.

#### Task performance

At baseline, 10 patients lacked behavioral task data (*N*=341; 4 missing data; 7 poor performance). At two-year follow-up, 34 patients (*N*=291; 24 missing data; 10 poor performance) and 2 controls (*N*=54; 2 missing data) lacked behavioral task data.

#### Functional MRI

At baseline, 30 patients lacked functional MRI data (*N*=332; 1 missing scan; 4 missing behavioral data; 13 technical issues; 3 poor performance; 9 poor image quality). At two-year follow-up, 53 patients (*N*=276; 28 missing scan; 10 missing behavioral data; 3 technical issues; 3 poor performance; 9 poor image quality) and 4 controls (*N*=52; 2 missing scan; 2 quality control) lacked functional MRI data.

#### Diffusion-weighted MRI

At baseline, 1 patients lacked diffusion-weighted MRI data (*N*=351; 1 missing scan). At two-year follow-up, 22 patients (*N*=307; 22 missing scan) and 2 controls (*N*=54; 2 missing scan) lacked diffusion-weighted data.

For analyses of substantia nigra free water, 19 patients were excluded at baseline (*N*=332; 7 technical issues, 12 perivascular spaces) and 22 patients were excluded at follow-up (*N*=285; 10 technical issues, 12 perivascular spaces). Additionally, 10 controls were excluded at baseline (*N*=50) and at follow-up (*N*=45) due to perivascular spaces.

For analyses of cortical mean diffusivity, 13 patients were excluded at baseline (*N*=338; 13 technical issues) and 11 patients were excluded at follow-up (*N*=296; 11 technical issues).

### Behavioral performance metrics pertaining to response strategies

PD-related behavioral deficits may prompt patients to adopt response strategies in the action selection task that differ from the response strategies of healthy controls. This may introduce additional variability into measurements of brain activity, thereby exerting confounding influences on our functional MRI analyses. To ascertain whether such a confounding influence may be present in our data, we investigated additional metrics of behavioral performance that more closely capture potential discrepancies in response strategies between PD patients and healthy controls. In addition, we examined whether changes in these behavioral performance metrics correlated with bradykinesia progression.

#### Error rates

PD leads to cognitive deficits and impaired coordination of individual finger movements, which may impair action selection capabilities. In the action selection task, this may manifest as increased error rates, resulting from responses that do not reflect intended targets. We therefore investigated whether PD differentially influenced longitudinal changes in error rates. We observed a general increase in error rates (logistic mixed-effects model, TIME effect: *χ*^2^(1)=8.75, *P*=0.003; follow-up>baseline: *OR*=1.56, *SE*=0.23), suggestive of an aging-related decline in accuracy. Alternatively, in light of our finding that response times decreased over time, this may indicate that participants favored speed over accuracy to a greater extent at follow-up compared to baseline. We also observed a trend towards higher error rates in PD patients compared to healthy controls (logistic mixed-effects model, GROUP effect: *χ*^2^(1)=3.29, *P*=0.07; patient>control: *OR*=1.43, *SE*=0.28), and a trend towards a general increase in error rates for multiple-choice compared to single-choice (logistic mixed-effects model, CHOICE effect *χ*^2^(1)=3.05, *P*=0.08; multiple>single: *OR*=1.24, *SE*=0.15).

We observed no relationship between longitudinal changes in error rates and bradykinesia progression (logistic mixed-effects model: *P*=0.47). Nevertheless, we observed that error rates correlated with the severity of bradykinesia. More specifically, bradykinesia severity correlated with motor-related error rates (multiple linear regression: *F*(1)=13.22, *P*<0.001, *β*=1.41e-03, *SE*=3.90e-04) and selection-related error rates (multiple linear regression: *F*(1)=9.218, *P*=0.003, *β*=-1.20e-03, *SE*=4.0e-04) at baseline. There were no correlations between error rates and bradykinesia at follow-up. These findings suggest that appropriate target selection may become more difficult as bradykinesia severity increases (multiple linear regression: *P*>0.13).

#### Missed responses

PD also impairs the ability to initiate actions, potentially resulting in more missed responses in the action selection task. We therefore investigated whether PD differentially influenced longitudinal changes in numbers of missed responses. We observed a longitudinal increase in misses, which was smaller for patients than controls (Poisson mixed-effects model, GROUP×TIME effect: *χ*^2^(1)=5.56, *P*=0.018; follow-up>baseline, patients>controls: *OR*=0.59, *SE*=0.13), although both groups missed more responses over time (follow-up>baseline, patients: *OR*=1.33, *SE*=0.08, *z-ratio*=4.62, *P*<0.001; follow-up>baseline, controls: *OR*=2.26, *SE*=0.49, *z-ratio*=3.77, *P*<0.001). Consistent with our analysis of error rates, these results point to aging-related effects on cognition, demonstrating that PD has limited effects on the ability to respond accurately and timely to the action selection task.

We observed no association between bradykinesia progression and longitudinal change in misses (multiple linear regression: *P*=0.68). Nevertheless, we observed that number of misses correlated with bradykinesia severity. More specifically, bradykinesia severity correlated with number of misses at both baseline (Poisson regression: *χ*^2^(1)=44.00, *P*<0.001, *IRR*=1.04, *SE*=0.006) and follow-up (Poisson regression: *χ*^2^(1)=51.45, *P*<0.001, *IRR*=1.05, *SE*=0.008).

#### Response flexibility

PD places constraints on the ability to flexibly adjust response strategies according to changes in the environment, manifesting as a tendency to repeat responses rather than selecting new ones. In this study, participants were asked to vary their responses as much as possible during the action selection task, enabling us to test whether PD is characterized by reduced behavioral flexibility, and how this reduced flexibility may relate to clinical progression, which we investigated in two ways.

##### Response switching

First, we examined whether PD patients were more prone to repeating responses in trials where they had the option to switch to a new one. We quantified response switching ability as the ratio between numbers of response repeats and response switches, accounting only for trials in which both repeats and switches were simultaneously possible. We then tested whether PD differentially influenced longitudinal changes in response switching. We observed a trend towards lower switching rates in PD patients compared to healthy controls (linear mixed-effects model, GROUP effect: *χ*^2^(1)=3.69, *P*=0.055; patient>control: *log-ratio*=0.90, *SE*=0.05). These findings indicate that PD patients may be more prone to repetitive responding strategies.

We also observed that faster bradykinesia progression was associated with longitudinal decreases in switching rates (multiple linear regression: *F*(1)=4.38, *P*=0.037, *β*=-0.41, *SE*=0.19), suggesting that responding strategies become more repetitive as symptoms worsen. However, there were no correlations between bradykinesia at either baseline (multiple linear regression: *P*>0.9) or follow-up (multiple linear regression: *P*>0.18).

##### Response variability

Second, we examined whether PD patients had reduced variability in selecting responses throughout the action selection task. We quantified response variability as the coefficient of variation among the four possible response options. We then investigated whether PD differentially influenced longitudinal changes in response variability. We observed that PD patients had higher response variability compared to healthy controls (linear mixed-effects model, GROUP effect: *χ*^2^(1)=4.49, *P*=0.034, *η*^2^ =0.012; patient>control: *log-ratio*=1.12, *SE*=0.10, *t-ratio*(380)=2.12, *P*=0.035). We also observed that response variability generally increased over time (linear mixed-effects model, TIME effect: *χ*^2^(1)=4.60, *P*=0.032, *η^2^_p_*=0.013; follow-up>baseline: *log-ratio*=1.13, *SE*=0.06, *t-ratio*(355)=2.14, *P*=0.033). These findings indicate that PD patients were capable of varying their responses during the action selection task. This eliminates the potential confound that PD patients and healthy controls differ in their reliance on habitual versus goal-directed action selection during the task.

We observed no association between bradykinesia progression and longitudinal change in response variability (multiple linear regression: *P*=0.37). Nevertheless, we observed that bradykinesia severity correlated with response variability at baseline (multiple linear regression: *F*(1)=5.12, *P*=0.024, *β*=3.18e-03, *SE*=1.41e-03), but not at follow-up (multiple linear regression: *P*=0.39).

### Missingness of data

At baseline, 351 patients and 60 controls participated. 329 patients and 56 controls returned for two-year follow-up measurements.

#### Clinical data

At baseline, 23 ON-state (*N*=329) and 6 OFF-state (*N*=346) patients lacked clinical data. At two-year follow-up, 14 ON-state (*N*=315) and 18 OFF-state (*N*=311) patients lacked clinical data.

#### Task performance

At baseline, 10 patients lacked behavioral task data (*N*=341; 4 missing data; 7 poor performance). At two-year follow-up, 34 patients (*N*=291; 24 missing data; 10 poor performance) and 2 controls (*N*=54; 2 missing data) lacked behavioral task data.

#### Functional MRI

At baseline, 30 patients lacked functional MRI data (*N*=332; 1 missing scan; 4 missing behavioral data; 13 technical issues; 3 poor performance; 9 poor image quality). At two-year follow-up, 53 patients (*N*=276; 28 missing scan; 10 missing behavioral data; 3 technical issues; 3 poor performance; 9 poor image quality) and 4 controls (*N*=52; 2 missing scan; 2 quality control) lacked functional MRI data.

#### Diffusion-weighted MRI

At baseline, 1 patients lacked diffusion-weighted MRI data (*N*=351; 1 missing scan). At two-year follow-up, 22 patients (*N*=307; 22 missing scan) and 2 controls (*N*=54; 2 missing scan) lacked diffusion-weighted data.

For analyses of substantia nigra free water, 19 patients were excluded at baseline (*N*=332; 7 technical issues, 12 perivascular spaces) and 22 patients were excluded at follow-up (*N*=285; 10 technical issues, 12 perivascular spaces). Additionally, 10 controls were excluded at baseline (*N*=50) and at follow-up (*N*=45) due to perivascular spaces.

For analyses of cortical mean diffusivity, 13 patients were excluded at baseline (*N*=338; 13 technical issues) and 11 patients were excluded at follow-up (*N*=296; 11 technical issues).

### Behavioral performance metrics pertaining to response strategies

PD-related behavioral deficits may prompt patients to adopt response strategies in the action selection task that differ from the response strategies of healthy controls. This may introduce additional variability into measurements of brain activity, thereby exerting confounding influences on our functional MRI analyses. To ascertain whether such a confounding influence may be present in our data, we investigated additional metrics of behavioral performance that more closely capture potential discrepancies in response strategies between PD patients and healthy controls. In addition, we examined whether changes in these behavioral performance metrics correlated with bradykinesia progression.

#### Error rates

PD leads to cognitive deficits and impaired coordination of individual finger movements, which may impair action selection capabilities. In the action selection task, this may manifest as increased error rates, resulting from responses that do not reflect intended targets. We therefore investigated whether PD differentially influenced longitudinal changes in error rates. We observed a general increase in error rates (logistic mixed-effects model, TIME effect: *χ*^2^(1)=8.75, *P*=0.003; follow-up>baseline: *OR*=1.56, *SE*=0.23), suggestive of an aging-related decline in accuracy. Alternatively, in light of our finding that response times decreased over time, this may indicate that participants favored speed over accuracy to a greater extent at follow-up compared to baseline. We also observed a trend towards higher error rates in PD patients compared to healthy controls (logistic mixed-effects model, GROUP effect: *χ*^2^(1)=3.29, *P*=0.07; patient>control: *OR*=1.43, *SE*=0.28), and a trend towards a general increase in error rates for multiple-choice compared to single-choice (logistic mixed-effects model, CHOICE effect *χ*^2^(1)=3.05, *P*=0.08; multiple>single: *OR*=1.24, *SE*=0.15).

We observed no relationship between longitudinal changes in error rates and bradykinesia progression (logistic mixed-effects model: *P*=0.47). Nevertheless, we observed that error rates correlated with the severity of bradykinesia. More specifically, bradykinesia severity correlated with motor-related error rates (multiple linear regression: *F*(1)=13.22, *P*<0.001, *β*=1.41e-03, *SE*=3.90e-04) and selection-related error rates (multiple linear regression: *F*(1)=9.218, *P*=0.003, *β*=-1.20e-03, *SE*=4.0e-04) at baseline. There were no correlations between error rates and bradykinesia at follow-up. These findings suggest that appropriate target selection may become more difficult as bradykinesia severity increases (multiple linear regression: *P*>0.13).

#### Missed responses

PD also impairs the ability to initiate actions, potentially resulting in more missed responses in the action selection task. We therefore investigated whether PD differentially influenced longitudinal changes in numbers of missed responses. We observed a longitudinal increase in misses, which was smaller for patients than controls (Poisson mixed-effects model, GROUP×TIME effect: *χ*^2^(1)=5.56, *P*=0.018; follow-up>baseline, patients>controls: *OR*=0.59, *SE*=0.13), although both groups missed more responses over time (follow-up>baseline, patients: *OR*=1.33, *SE*=0.08, *z-ratio*=4.62, *P*<0.001; follow-up>baseline, controls: *OR*=2.26, *SE*=0.49, *z-ratio*=3.77, *P*<0.001). Consistent with our analysis of error rates, these results point to aging-related effects on cognition, demonstrating that PD has limited effects on the ability to respond accurately and timely to the action selection task.

We observed no association between bradykinesia progression and longitudinal change in misses (multiple linear regression: *P*=0.68). Nevertheless, we observed that number of misses correlated with bradykinesia severity. More specifically, bradykinesia severity correlated with number of misses at both baseline (Poisson regression: *χ*^2^(1)=44.00, *P*<0.001, *IRR*=1.04, *SE*=0.006) and follow-up (Poisson regression: *χ*^2^(1)=51.45, *P*<0.001, *IRR*=1.05, *SE*=0.008).

#### Response flexibility

PD places constraints on the ability to flexibly adjust response strategies according to changes in the environment, manifesting as a tendency to repeat responses rather than selecting new ones. In this study, participants were asked to vary their responses as much as possible during the action selection task, enabling us to test whether PD is characterized by reduced behavioral flexibility, and how this reduced flexibility may relate to clinical progression, which we investigated in two ways.

##### Response switching

First, we examined whether PD patients were more prone to repeating responses in trials where they had the option to switch to a new one. We quantified response switching ability as the ratio between numbers of response repeats and response switches, accounting only for trials in which both repeats and switches were simultaneously possible. We then tested whether PD differentially influenced longitudinal changes in response switching. We observed a trend towards lower switching rates in PD patients compared to healthy controls (linear mixed-effects model, GROUP effect: *χ*^2^(1)=3.69, *P*=0.055; patient>control: *log-ratio*=0.90, *SE*=0.05). These findings indicate that PD patients may be more prone to repetitive responding strategies.

We also observed that faster bradykinesia progression was associated with longitudinal decreases in switching rates (multiple linear regression: *F*(1)=4.38, *P*=0.037, *β*=-0.41, *SE*=0.19), suggesting that responding strategies become more repetitive as symptoms worsen. However, there were no correlations between bradykinesia at either baseline (multiple linear regression: *P*>0.9) or follow-up (multiple linear regression: *P*>0.18).

##### Response variability

Second, we examined whether PD patients had reduced variability in selecting responses throughout the action selection task. We quantified response variability as the coefficient of variation among the four possible response options. We then investigated whether PD differentially influenced longitudinal changes in response variability. We observed that PD patients had higher response variability compared to healthy controls (linear mixed-effects model, GROUP effect: *χ*^2^(1)=4.49, *P*=0.034, *η*^2^ =0.012; patient>control: *log-ratio*=1.12, *SE*=0.10, *t-ratio*(380)=2.12, *P*=0.035). We also observed that response variability generally increased over time (linear mixed-effects model, TIME effect: *χ*^2^(1)=4.60, *P*=0.032, *η^2^_p_*=0.013; follow-up>baseline: *log-ratio*=1.13, *SE*=0.06, *t-ratio*(355)=2.14, *P*=0.033). These findings indicate that PD patients were capable of varying their responses during the action selection task. This eliminates the potential confound that PD patients and healthy controls differ in their reliance on habitual versus goal-directed action selection during the task.

We observed no association between bradykinesia progression and longitudinal change in response variability (multiple linear regression: *P*=0.37). Nevertheless, we observed that bradykinesia severity correlated with response variability at baseline (multiple linear regression: *F*(1)=5.12, *P*=0.024, *β*=3.18e-03, *SE*=1.41e-03), but not at follow-up (multiple linear regression: *P*=0.39).

**Supplementary Table 1.** Group analyses of longitudinal changes in brain structure.

| Area | Healthy | PD | GROUP×TIME<br>($\chi^2$) | Effect size<br>($\eta^2_p$) |
| --- | --- | --- | --- | --- |
| ***ΔSubstantia nigra free water*** |  |  |  |  |
| Anterior | 0.998 (0.042) | 0.973 (0.016) | 0.332 | - |
| Posterior | 0.990 (0.020) | 1.027 (0.008)*** | 2.946+ | 0.031 |
| ***ΔCortical mean diffusivity (un-corrected)*** |  |  |  |  |
| Caudal middle frontal | 1.009 (0.003)*** | 1.016 (0.001)*** | 5.523* | 0.015 |
| Inferior parietal | 1.009 (0.003)*** | 1.020 (0.001)*** | 14.377*** | 0.039 |
| Paracentral | 1.006 (0.004) | 1.022 (0.002)*** | 12.867** | 0.035 |
| Postcentral | 1.011 (0.003)*** | 1.017 (0.001)*** | 3.596 | - |
| Precentral | 1.015 (0.003)*** | 1.015 (0.001)*** | 0.025 | - |
| Precuneus | 1.002 (0.003) | 1.017 (0.001)*** | 22.650*** | 0.060 |
| Superior frontal | 1.008 (0.002)*** | 1.016 (0.001)*** | 8.357** | 0.023 |
| Superior parietal | 1.015 (0.003)*** | 1.019 (0.001)*** | 1.748 | - |
| Supramarginal | 1.010 (0.003)*** | 1.020 (0.001)*** | 11.067** | 0.031 |
| ***ΔCortical mean diffusivity (free water-corrected)*** |  |  |  |  |
| Caudal middle frontal | 1.000 (0.001) | 1.003 (0.001)*** | 6.717* | 0.019 |
| Inferior parietal | 1.000 (0.002) | 1.001 (0.001)+ | 0.219 | - |
| Paracentral | 1.001 (0.001) | 1.005 (0.001)*** | 9.332* | 0.026 |
| Postcentral | 1.005 (0.001)*** | 1.003 (0.001)*** | 1.156 | - |
| Precentral | 1.005 (0.001)*** | 1.003 (0.001)*** | 1.689 | - |
| Precuneus | 0.998 (0.001)+ | 1.002 (0.000)*** | 16.046*** | 0.044 |
| Superior frontal | 1.001 (0.001) | 1.004 (0.000)*** | 6.443* | 0.018 |
| Superior parietal | 1.003 (0.001)** | 1.003 (0.000)*** | 0.186 | - |
| Supramarginal | 1.002 (0.001) | 1.003 (0.001)*** | 0.155 | - |

Values for each group are displayed as log-ratios (standard errors) of differences between sessions (follow-up – baseline) extracted from linear mixed-effects models using the *emmeans* package in R. P-values for GROUP×TIME interactions in cortical MD analyses were adjusted for multiple comparisons using the false-discovery rate method (n=9). Δ=Follow-up>Baseline, +=P<0.1, \*=P<0.05, \*\*=P<0.01, \*\*\*=P<0.001.

**Supplementary table 2.**
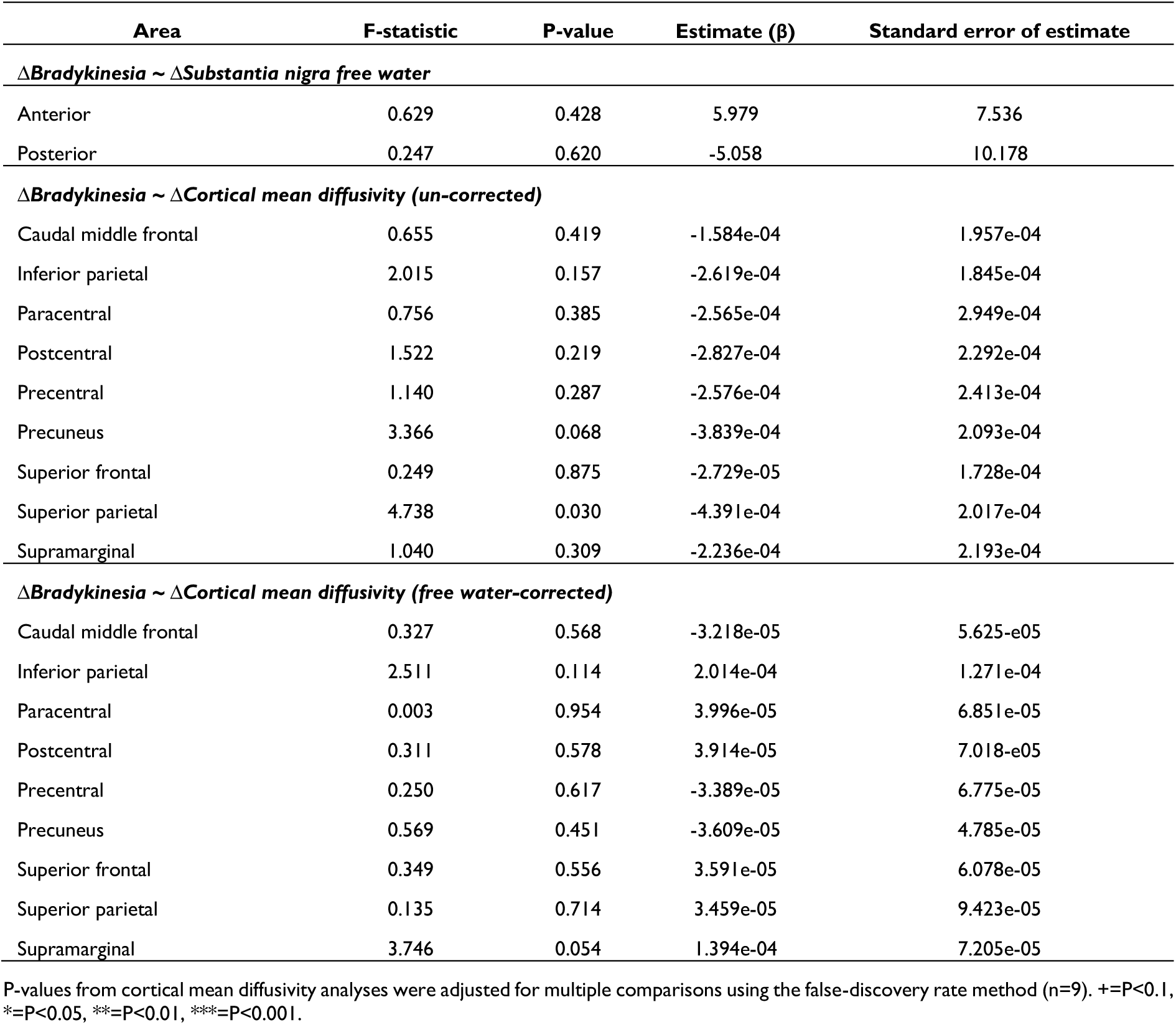
Correlations between bradykinesia progression and longitudinal changes in structural MRI-based metrics.

**Supplementary table 3.** Correlations between bradykinesia severity and structural MRI-based metrics at baseline.

| Area | F-statistic | P-value | Estimate (β) | Standard error of estimate |
| --- | --- | --- | --- | --- |
| ***Bradykinesia severity ~ Substantia nigra free water*** |  |  |  |  |
| Anterior | 2.837 | 0.093 | 3.543e-03 | 2.103e-03 |
| Posterior | 2.659 | 0.104 | -2.576e-03 | 1.580e-03 |
| ***Bradykinesia severity ~ Cortical mean diffusivity (un-corrected)*** |  |  |  |  |
| Caudal middle frontal | 5.207 | 0.0232 | 5.772e-04 | 2.530e-04 |
| Inferior parietal | 6.985 | 8.635e-03* | 7.659e-04 | 2.898e-04 |
| Paracentral | 1.001 | 0.318 | 3.255e-04 | 3.253e-04 |
| Postcentral | 3.147 | 0.077 | 5.863e-04 | 3.305e-04 |
| Precentral | 2.661 | 0.104 | 5.178e-04 | 3.174e-04 |
| Precuneus | 4.278 | 0.0394* | 5.916e-04 | 2.860e-04 |
| Superior frontal | 5.358 | 0.0212* | 5.226e-04 | 2.258e-04 |
| Superior parietal | 0.576 | 0.449 | 2.741e-04 | 3.612e-04 |
| Supramarginal | 7.708 | 5.832e-03* | 7.711e-04 | 2.778e-04 |
| ***Bradykinesia severity ~ Cortical mean diffusivity (free water-corrected)*** |  |  |  |  |
| Caudal middle frontal | 1.129 | 0.289 | 1.108e-04 | 1.043e-04 |
| Inferior parietal | 1.288 | 0.257 | 1.116e-04 | 9.831e-05 |
| Paracentral | 0.495 | 0.482 | 8.744e-05 | 1.243e-04 |
| Postcentral | 0.832 | 0.362 | 1.053e-04 | 1.154e-04 |
| Precentral | 1.155 | 0.283 | 1.389e-04 | 1.292e-04 |
| Precuneus | 2.771 | 0.097 | 1.331e-04 | 7.997e-05 |
| Superior frontal | 1.600 | 0.207 | 1.096e-04 | 8.662e-05 |
| Superior parietal | 0.889 | 0.347 | 9.986e-05 | 1.059e-04 |
| Supramarginal | 1.608 | 0.258 | 1.169e-04 | 9.218e-05 |

P-values from cortical mean diffusivity analyses were adjusted for multiple comparisons using the false-discovery rate method (n=9). +=P<0.1, \*=P<0.05, \*\*=P<0.01, \*\*\*=P<0.001.

## Notes

### Clinical Protocols

https://link.springer.com/article/10.1186/s12883-019-1394-3

### Author Declarations

The study was approved by a medical ethical committee (METC Oost-Nederland, formerly CMO Arnhem-Nijmegen; #2016-2934 and #2018-4785).

